# A Simple Mathematical Tool to Help Distribute Doses of ‘Two-Dose’ Covid-19 Vaccines among Non-Immunized and Partly-Immunized Population

**DOI:** 10.1101/2021.05.10.21256978

**Authors:** Aanandita Kapoor, Krishan Mohan Kapoor

## Abstract

**Background:** Full immunization with two doses of Covid vaccine has been found to be a critical factor in preventing morbidity and mortality from the Covid-19 infection. However, due to the shortage of vaccines, a significant portion of the population is not getting vaccination in many countries. Also, the distribution of vaccine doses between prospective first dose recipient and second dose recipient is not uniformly planned, as seen in India’s various states and union territories. It is recommended to give second vaccine doses within 4-8 weeks to first dose recipients for both the approved vaccines in India; hence the judicious distribution between non-immunized and partly immunized populations is essential. Managing the Covid-19 vaccination drive in an area with a large number of single-dose recipients compared to a smaller number of fully immunized people can become a huge administrative challenge. Therefore, this study was conducted to assess the number of people covered under the Covid vaccination drive in India and analyze the state-wise distribution of vaccines among the non-immunized and partly immunized population.

**Methods:** The Covid 19 vaccination data till 7^th^ may, 2021 was taken from the website of the Ministry of Health and Family Welfare, Govt of India. From the data available of the number of doses injected, other figures like the total number of people vaccinated, people with two doses of vaccine or full immunization (FI), and those with a single dose of vaccine or partial immunization (PI) were found. The percentage of the fully immunized and partly immunized population was also found. A ratio between fully immunized and partly immunized individuals (FI: PI) was proposed as a guide to monitor the progress of the vaccination and future dose distribution of ‘two-dose’ Covid-19 vaccines among partly immunized (PI) and non-immunized (NI) population.

**Results:** In India, till 7 May 2021, 16,49,73,058 doses of Covid-19 vaccines have been injected. A total of 13,20,87,824 people received these vaccine doses, with 9,92,02,590 people getting a single dose or were partly immunized (PI), and 3,28,85,234 got two doses each or were fully immunized (FI). Among the states, Tripura and Andhra Pradesh had the highest FI: PI (Fully Immunized: Partly Immunized) ratio of 0.86 and 0.52, followed by Tamil Nadu, Arunachal Pradesh, and West Bengal with figures of 0.48. 0.47 and 0.47, respectively. Telangana and Punjab had the lowest FI: PI ratio among the states at 0.2 each, with Chhattisgarh, Madhya Pradesh, and Haryana following at 0.21. 0.23 and 0.23, respectively. These values are much lower than the national average of 0.33 in India.

**Conclusion:** The FI: PI ratio could help governments decide how to use scarce vaccine resources among first-time and second-time recipients. This simple mathematical tool could ensure full immunization status to maximum people within the recommended 4-8 week time window after the first dose to avoid a large population group with partly immunized status.

## Introduction

The covid vaccine has been found to be a critical factor in preventing morbidity and mortality from the Covid-19 infection^1 2 3^. Globally, the governments are trying to immunize their people as fast as possible to achieve maximum population coverage. For the Covid-19 vaccination drive to be effective, three things need to be planned: supply of vaccines, people needed to implement vaccination, and distribution among people to be vaccinated. If there is a mismatch in these factors, failure of the vaccination drive is inevitable.^4^ As the pace of the COVID-19 campaign in India is being driven by vaccine supplies, distribution between non-immunized and partly immunized people assumes significance for the success of the Covid-19 vaccination drive. It is recommended to give a second dose of the Covid-19 vaccine within 4-8 weeks of the first dose, so it is crucial to distribute the vaccine doses among the non-immunized and partly immunized population. Having many more partly immunized people than fully immunized people in a geographical area can lead to many problems in the vaccination drive against Covid-19. This study was conducted on Indian vaccination data to find a simple mathematical tool that could help distribute Covid-19 vaccine doses among the non-immunized and partly immunized population.

## Material and Methods

The national and state & union territories data of Covid 19 vaccination in India till 7 am of 7^th^ may, 2021 was taken from the official website of Ministry of Health and Family Welfare, Govt of India. (https://www.mohfw.gov.in/pdf/CumulativeCOVIDVaccinationCoverageReport06052021.pdf) The data gets updated every day. The data presented the national and state-wise total number of doses injected, the total number of single doses injected, and a total number of second doses injected. From the data available about the number of doses injected, other figures like the total number of people vaccinated, people with two doses of vaccine or full immunization (FI), and people with one dose of vaccine or partial immunization (PI) were found. The percentage of fully immunized and partly immunized was also calculated among the vaccine beneficiaries. A ratio between fully immunized and partly immunized individuals (FI: PI) was calculated and proposed as a simple guide to monitoring the progress of the vaccination and dose distribution of ‘two-dose’ Covid-19 vaccines among partly immunized (PI) and non-immunized (NI) population. No ethics board approval was needed for this research paper, as this information was available in the public domain.

## Results

In India, till 7 May, 16,49,73,058 doses of Covid-19 vaccines have been injected. A total of 13,20,87,824 people received these vaccine doses, with 9,92,02,590 people getting a single dose or were partly immunized (PI), as shown in Table I.

**Table I:**
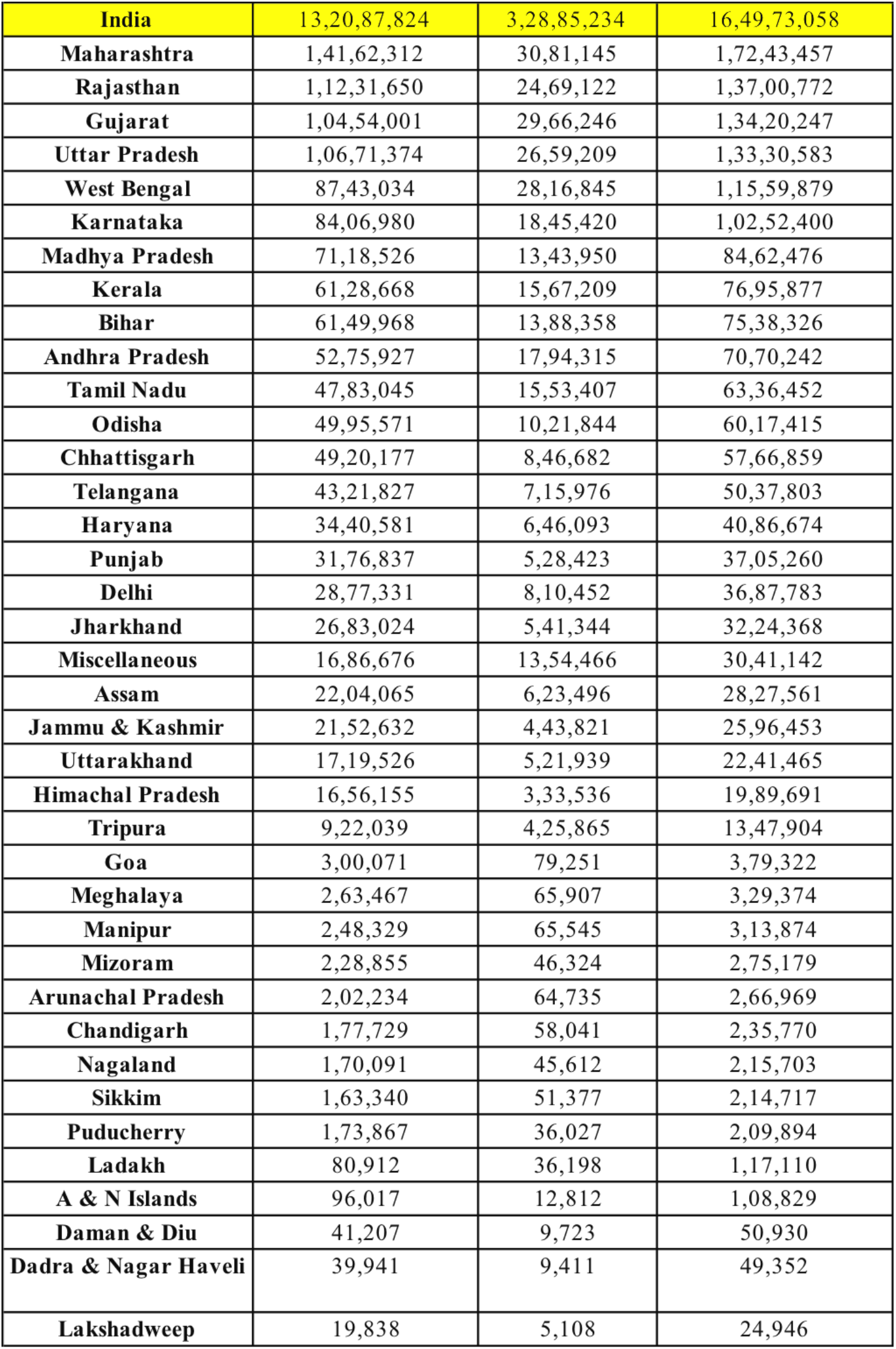
Showing the total number of doses, first doses, and second doses injected in various states and Union Territories of India.

A total of 3,28,85,234 people got two doses each or were fully immunized (FI). The population not covered under any of these groups was categorized as non-immunized (NI). The same data was worked out for every state and union territory as given in Table II.

**Table II:** Showing the total number of recipients, first dose beneficiaries or partly Immunized (PI), 2^nd^ doses recipients or Fully immunized (FI), and the FI: PI ratio in various states & UT’s of India.

| State/UT | Partly Immunized with Single dose (PI) | PI % age | Fully Immunized with Two Doses (FI) | FI %age | Total Vaccine Beneficiaries | FI:PI Ratio |
| --- | --- | --- | --- | --- | --- | --- |
| Miscellaneous | 3,32,210 | 19.7 | 13,54,466 | 80.3 | 16,86,676 | 4.07 |
| Tripura | 4,96,174 | 53.8 | 4,25,865 | 46.2 | 9,22,039 | 0.86 |
| Ladakh | 44,714 | 55.3 | 36,198 | 44.7 | 80,912 | 0.81 |
| Andhra Pradesh | 34,81,612 | 66 | 17,94,315 | 34 | 52,75,927 | 0.52 |
| Chandigarh | 1,19,688 | 67.3 | 58,041 | 32.7 | 1,77,729 | 0.48 |
| Tamil Nadu | 32,29,638 | 67.5 | 15,53,407 | 32.5 | 47,83,045 | 0.48 |
| Arunachal Pradesh | 1,37,499 | 68 | 64,735 | 32 | 2,02,234 | 0.47 |
| West Bengal | 59,26,189 | 67.8 | 28,16,845 | 32.2 | 87,43,034 | 0.47 |
| Sikkim | 1,11,963 | 68.5 | 51,377 | 31.5 | 1,63,340 | 0.46 |
| Uttarakhand | 11,97,587 | 69.6 | 5,21,939 | 30.4 | 17,19,526 | 0.44 |
| Gujarat | 74,87,755 | 71.6 | 29,66,246 | 28.4 | 1,04,54,001 | 0.4 |
| Assam | 15,80,569 | 72 | 6,23,496 | 28 | 22,04,065 | 0.39 |
| Delhi | 20,66,879 | 71.8 | 8,10,452 | 28.2 | 28,77,331 | 0.39 |
| Nagaland | 1,24,479 | 72.2 | 45,612 | 27.8 | 1,70,091 | 0.37 |
| Goa | 2,20,820 | 73.6 | 79,251 | 26.4 | 3,00,071 | 0.36 |
| Manipur | 1,82,784 | 73.6 | 65,545 | 26.4 | 2,48,329 | 0.36 |
| Lakshadweep | 14,730 | 74.3 | 5,108 | 25.7 | 19,838 | 0.35 |
| Kerala | 45,61,459 | 74.4 | 15,67,209 | 25.6 | 61,28,668 | 0.34 |
| Meghalaya | 1,97,560 | 75 | 65,907 | 25 | 2,63,467 | 0.33 |
| Uttar Pradesh | 80,12,165 | 75.1 | 26,59,209 | 24.9 | 1,06,71,374 | 0.33 |
| **India** | **9,92,02,590** | **75.1** | **32885234** | **24.9** | **13,20,87,824** | **0.33** |
| Dadra & Nagar Haveli | 30,530 | 76.4 | 9,411 | 23.6 | 39,941 | 0.31 |
| Daman & Diu | 31,484 | 76.4 | 9,723 | 23.6 | 41,207 | 0.31 |
| Bihar | 47,61,610 | 77.4 | 13,88,358 | 22.6 | 61,49,968 | 0.29 |
| Karnataka | 65,61,560 | 78 | 18,45,420 | 22 | 84,06,980 | 0.28 |
| Maharashtra | 1,10,81,167 | 78.2 | 30,81,145 | 21.8 | 1,41,62,312 | 0.28 |
| Rajasthan | 87,62,528 | 78 | 24,69,122 | 22 | 1,12,31,650 | 0.28 |
| Jammu & Kashmir | 17,08,811 | 79.4 | 4,43,821 | 20.6 | 21,52,632 | 0.26 |
| Odisha | 39,73,727 | 79.5 | 10,21,844 | 20.5 | 49,95,571 | 0.26 |
| Puducherry | 1,37,840 | 79.3 | 36,027 | 20.7 | 1,73,867 | 0.26 |
| Himachal Pradesh | 13,22,619 | 79.9 | 3,33,536 | 20.1 | 16,56,155 | 0.25 |
| Jharkhand | 21,41,680 | 79.8 | 5,41,344 | 20.2 | 26,83,024 | 0.25 |
| Mizoram | 1,82,531 | 79.6 | 46,324 | 20.4 | 2,28,855 | 0.25 |
| Haryana | 27,94,488 | 81.2 | 6,46,093 | 18.8 | 34,40,581 | 0.23 |
| Madhya Pradesh | 57,74,576 | 81.1 | 13,43,950 | 18.9 | 71,18,526 | 0.23 |
| Chhattisgarh | 40,73,495 | 82.2 | 8,46,682 | 17.8 | 49,20,177 | 0.21 |
| Punjab | 26,48,414 | 83.4 | 5,28,423 | 16.6 | 31,76,837 | 0.2 |
| Telangana | 36,05,851 | 83.4 | 7,15,976 | 16.6 | 43,21,827 | 0.2 |
| A & N Islands | 83,205 | 86.7 | 12,812 | 13.3 | 96,017 | 0.15 |

Maharashtra, Rajasthan, Uttar Pradesh, Gujarat, and West Bengal (in the decreasing order) had the highest number of total vaccine recipients in India while Ladakh, A & N Islands, Daman & Diu, Dadra & Nagar Haveli, and Lakshadweep (in the decreasing order) had the least number of vaccine recipient. (Figure 1)

**Figure 1:**
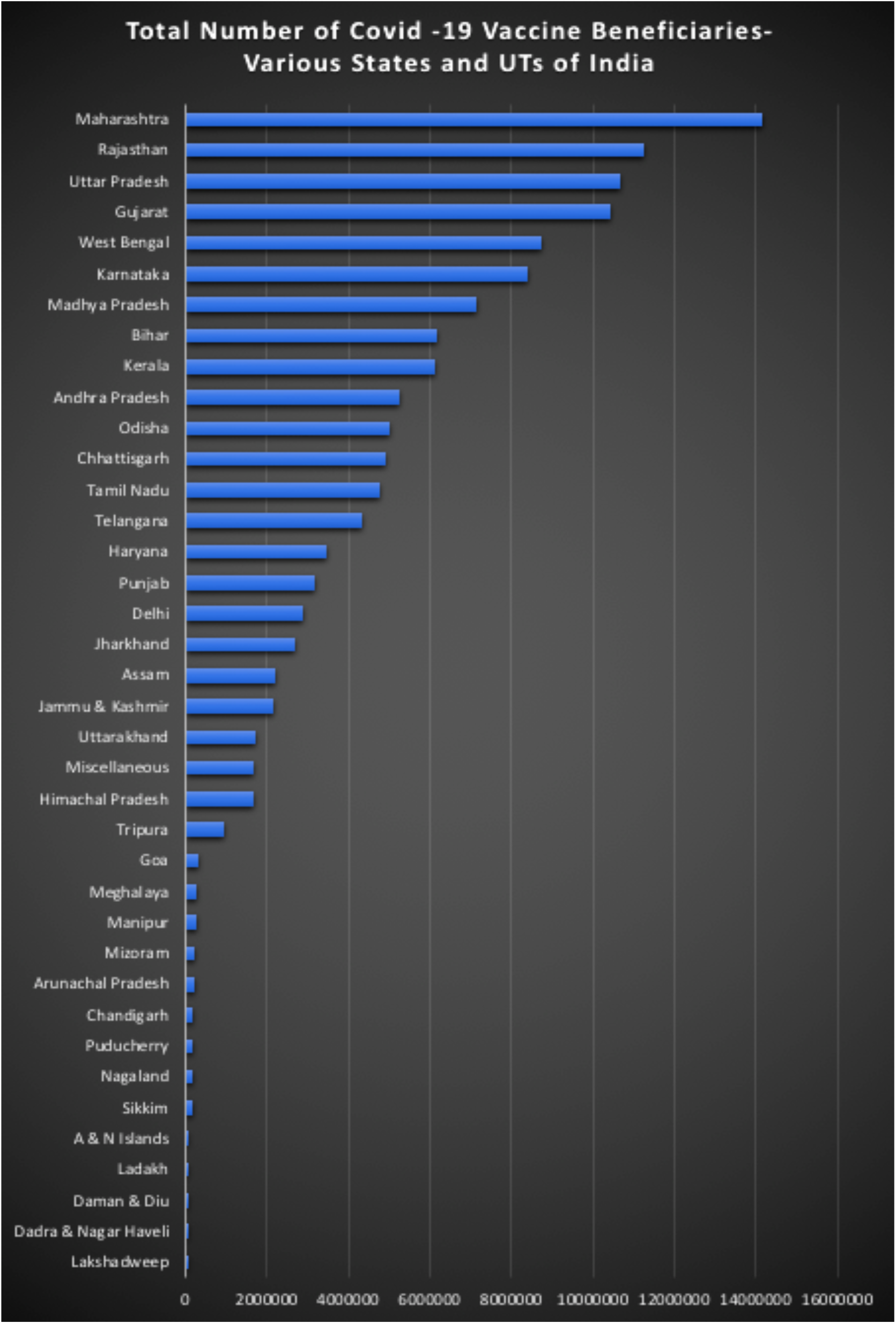
Showing the number of beneficiaries in various states & UTs of India, in descending order.

Among the states, Tripura and Andhra Pradesh had the highest FI: PI (fully immunized/ partly immunized) ratio of 0.86 and 0.52, followed by Tamil Nadu, Arunachal Pradesh, and West Bengal with figures of 0.48. 0.47 and 0.47, respectively. These values were much higher than the national average of 0.33 in India. Among the Union Territories, Ladakh and Chandigarh had the highest FI: PI ratio of and 0.48. (Figure 2)

**Figure 2:**
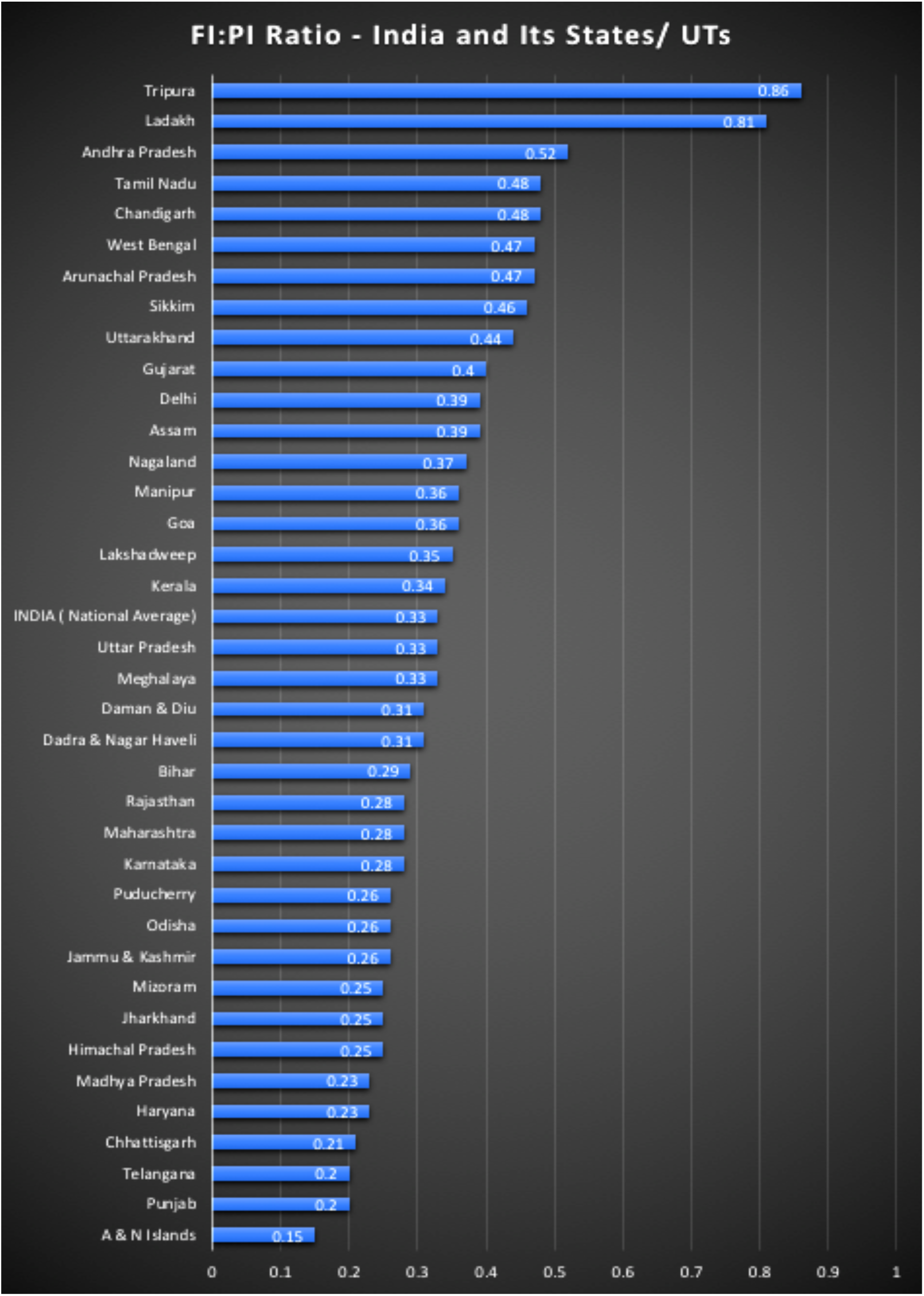
Showing FI: PI ratio in Various states and Union Territories of India (in descending order) * ‘Miscellaneous’ category not included in the graph.

Telangana and Punjab had the lowest FI: PI ratio among the states at 0.2 each, with Chhattisgarh, Madhya Pradesh, and Haryana at 0.21. 0.23 and 0.23, respectively. These values are much lower than the national average of 0.33 in India. Among the Union Territories, Andaman and Nicobar Islands had the lowest FI: PI ratio at 0.15, while Puducherry and Jammy & Kashmir had a low ratio of 0.26 each. The Miscellaneous group had the highest FI: PI ratio of 4.07 value.

## Discussion

The Covid -19 pandemic prompted scientists across the globe to design possible alternative COVID-19 vaccines^5^. Full immunization with two doses of Covid vaccine has been found to be a critical factor in preventing morbidity and mortality from the Covid-19 infection^1^. Globally the governments are trying to immunize their people as fast as possible to achieve maximum population coverage. The best strategy is to quickly deliver vaccines to as large a part of the population as possible and identify and tackle vaccine hesitancy sources^6^. This strategy would maximize the individual and population benefits of the approved COVID-19 vaccines.^7^

However, due to the shortage of vaccines in some countries, a significant number of the population is not getting access to vaccination^8^. Also, the vaccine distribution between prospective first dose recipient and second dose recipient is not uniformly planned in different geographic regions of the same country as seen in India’s various states and union territories. It is recommended to give a second vaccine dose within 4-8 weeks to first dose recipients for both the vaccines approved in India; hence distribution between non-immunized and partly immunized populations has become a challenge due to the limited time available. This present study was conducted to assess the number of people covered under the Covid vaccination drive in India and analyze the state-wise distribution of vaccines among non-immunized and partly immunized populations.

In India, the administration of COVID-19 vaccines started on 16 January 2021. As of 7 May 2021, India has administered 16,49,73,058 doses overall, including first & second doses of the currently approved vaccines. Two vaccines received approval for emergency use in India at the beginning of the vaccination program, i.e., Covishield ^9 10^ (the Oxford–AstraZeneca vaccine manufactured by the Serum Institute of India) and Covaxin (developed by an Indian company, Bharat Biotech).^11 12^ In April 2021, Sputnik V from Russia was also approved as a third vaccine, with its use expected to begin by late May 2021.

As per the Centre for Disease Control and Prevention (CDC), immunization is ‘a process by which a person becomes protected against a disease through vaccination.’ This term is frequently used interchangeably with vaccination. In this study, people who received two doses of the vaccine were called fully immunized, and those with only one vaccine dose were called partial immunized (PI) to calculate the FI: PI ratio.

The national average of the FI: PI value in India is 0.33, while there are states in India like Andhra Pradesh, Tamil Nadu, Arunachal Pradesh, and West Bengal with the FI: PI ratios of 0.52, 0.48. 0.47 and 0.47, respectively. These values are closer to the FI: PI values of 0.5. At the FI: PI value of 0.5, the planners in that area have given 50% of their total doses for full immunization of the same set of people by giving two doses (Fully Immunised or FI) while the remaining 50% doses were used as the first dose for another set of the population (partly Immunised or PI). It would now become straightforward for the planners to divide subsequent consignments equally in partly immunized and non-immunized populations.

It can also be explained like this; keeping the FI: PI ratio at 0.5 means that out of the first 100 vaccine doses, 25 persons have got two doses each & are fully immunized, and 50 persons have a single dose; hence they are partly immunized. The following 100 doses can be distributed ‘equally’ between 50 partly immunized (PI) recipients, waiting for their second dose, and 50 recipients from non-immunized (NI) population, getting their first dose. Similarly, subsequent batches can be distributed to keep a balance between fully immunized, partly immunized, and non-immunized populations. This process can be then continued indefinitely till the target population is vaccinated.

We can look at the case of Andhra Pradesh state with a FI: PI ratio of 0.52. Till 7^th^ May 2021 there were 34,81,612 single-dose recipients and 17,94,315 two-dose recipients (receiving 35,88,630 vaccine doses). This information shows great planning by the administrators handling the vaccine distribution work. Compare it with its neighboring Telangana state with a FI: PI ratio of 0.2 only. Till 7 May 2021, Telangana had a similar number of 36,05,851 single-dose recipients; however two-dose recipients dropped to only 7,15,976 (receiving only 14,31,952 vaccine doses). States with lower FI: PI scores have a huge task at their hand as they have to work very hard to get partly immunized people back for their second shots in the shortest possible time. At the same time, they will have to divert a large percentage of future vaccine consignment for partly immunized people; the scope of enrolling more non-immunized people for vaccination would be hit hard.

The danger of having a low FI: PI ratio in a geographic area is that there would be many more people with partial immunized than fully immunized people. Continuing to inject the first dose recipient may lead to a situation where the second dose recipient would miss out on their dose in 4-8 weeks. The gains associated with enrolling people for the first vaccine dose are also partly lost if the gap becomes very long before the second dose. This situation poses the following possible challenges in the vaccination drive against Covid-19.^13^

- There is a concern that people with only one dose might believe that they have adequate protection against COVID-19 infection, and they do not need the second dose. There is no evidence to show that people getting only one dose have adequate long-term protection against COVID-19 infection.^14^
- The potential risk from delaying the second dose of the Covid vaccine is that the second dose will be less effective when given later than the recommended interval.
- Also, many people who got their first vaccine may forget to return for their second dose after a long delay between the first and second dose.
- There is also a possibility that some people will be confused by regular changes in the vaccine schedule, and the resulting confusion may drive them to shun vaccination altogether or make them believe that they need a single dose only.
- Finally, some experts have warned that partial immunization with a single dose can lead to a less robust immune response giving only partial immunity against COVID-19 infection.
- Partial immunization may result in a greater risk that vaccine-resistant variants of SARS-CoV-2 could develop.

Many countries, including India, have a short time window available to take some decisive actions, and the vaccination drive could become a success story or a failure. Using a simple mathematical tool like FI: PI ratio for vaccine distribution is likely to make dose distribution between partly immunized and non-immunized people transparent and easily replicated across all the states.

## Limitations

1. The FI: PI ratio is not much relevant in single-dose Covid vaccines. It shows that there is a great need to develop single-dose vaccines to achieve fast population coverage.
2. In the absence of scarcity of doses and ample vaccine supply, the role of the FI: PI ratio is slightly less critical as coverage of non-immunized and partly immunized people can go on simultaneously. However, the FI: PI ratio still has a role in keeping a tract of partly immunized people to fully immunized ones.

## Conclusions

Vaccination coverage against Covid-19 in India needs much improvement at this point. The vaccine distribution in different states and union territories of the country is also not uniform. As a simple mathematical tool, the FI: PI ratio could help the governments decide how to use scarce Covid-19 vaccine resources among first-time and second-time recipients. This information can also help the government make changes in the vaccine booking app where the definitive assignment of slots among first-time and second-time recipients could control the FI: PI ratio. This method is likely to ensure full immunization status for maximum people within the prescribed 4-8 weeks after the first dose of Covid vaccine to avoid challenges posed by having a large population group with partly immunization status.

## Data Availability

The data referred to in the manuscript is available at the website of Ministry of Health and Family Welfare, India.

https://www.mohfw.gov.in/pdf/CumulativeCOVIDVaccinationCoverageReport06052021.pdf

## Abbreviations

FI: fully immunized
PI: partly immunized
NI: non-immunized

